# Human Respiratory Syncytial Virus Genetic Diversity and Lineage Replacement in Ireland pre- and post-COVID-19 pandemic

**DOI:** 10.1101/2024.07.23.24310850

**Authors:** Alan Rice, Gabriel Gonzalez, Michael Carr, Jonathan Dean, Emer O’Byrne, Lynn Aarts, Harry Vennema, Weronika Banka, Charlene Bennett, Siobhán Cleary, Lisa Domegan, Joan O’Donnell, Maureen O’Leary, Stephanie Goya, Lance Presser, Adam Meijer, Greg Martin, Hirofumi Sawa, Allison Waters, Cillian De Gascun, Daniel Hare

**Affiliations:** UCD National Virus Reference Laboratory, University College Dublin, Belfield, D04 E1W1, Ireland; Institute for Vaccine Research and Development, Hokkaido University, Hokkaido 001-0021, Japan; International Collaboration Unit, International Institute for Zoonosis Control, Hokkaido University, Sapporo, Hokkaido 001-0020, Japan; Centre for Infectious Disease Control, National Institute for Public Health and Environment, Bilthoven, the Netherlands; Health Protection Surveillance Centre, Dublin, Ireland; Department of Laboratory Medicine and Pathology, University of Washington, Seattle, USA; Health Improvement, Health Service Executive, Dublin, Ireland; Irish Blood Transfusion Service, National Blood Centre, Dublin, Ireland; UCD School of Public Health, Physiotherapy and Social Science, University College Dublin, Dublin, Ireland

**Keywords:** Human respiratory syncytial virus, whole-genome sequencing, genetic diversity, lineages, orthopneumovirus

## Abstract

**Background:** Human respiratory syncytial virus (HRSV) is a common cause of lower respiratory tract infections globally. Newly-licensed prophylactic vaccines and monoclonal antibodies are anticipated to alleviate this burden; however, such interventions may exert selective pressures on HRSV evolution.

**Methods:** Whole-genome sequencing was performed on HRSV-A (n=123) and -B (n=110) samples collected during three HRSV seasons in the 2021-2024 period from community cases. Additionally, G gene sequences, HRSV-A (n=141) and -B (n=141), collected in the 2015-2019 period were examined. Lineages were assigned by phylogenetic analyses including reference lineages.

**Results:** Phylogenetic trees inferred with the G gene and whole genomes were consistent. Changes in the prevalence of certain lineages post-COVID-19 reflected the impact of non-pharmaceutical interventions introduced to reduce SARS-CoV-2 transmission. The HRSV-A lineages A.D.1 and A.D.5 were dominant, while B.D.E.1 was the dominant lineage for HRSV-B. Similar trends were also observed in prevalent lineages in the European region. The emergence of a new lineage was identified as descended from A.D.1 with eight distinctive substitutions in proteins G, F and L. Other circulating lineages with amino acid substitutions were observed in the F glycoprotein which could impact binding sites of nirsevimab.

**Conclusion:** We provide the first comprehensive analysis of HRSV transmission and evolution in Ireland over the last decade through the selective forces created by the measures introduced during the COVID-19 pandemic. This study provides a foundation for future public health surveillance employing pathogen genomics to enable an evidence-based assessment of the impact of pharmaceutical interventions on HRSV evolution and disease severity.

**Key public health message:** *What did you want to address in this study and why?:* We aimed to address conditions enabling the yearly increase in the number of HRSV cases in recent years and the viral genetic diversity. A whole-genome sequencing-based molecular epidemiology of HRSV will be key to monitoring the effectiveness and impact of new immunisation programmes in the coming years.

*What have we learnt from this study?:* We have established a genomic-epidemiological baseline for HRSV in Ireland, and demonstrated a significant change in the diversity and abundance of viral lineages in circulation before, and after, the early years of the COVID-19 pandemic. Such changes in the most prevalent HRSV genetic lineages were shown to follow a similar trend across Europe during this time.

*What are the implications of your findings for public health?:* The characterised viral genetic diversity represents a benchmark for evidence-based future assessments of the effectiveness and the impact of new pharmaceutical interventions in Ireland i.e. monoclonal antibodies and HRSV vaccines for paediatric, geriatric and immunocompromised cohorts. Such preventive options are anticipated to reduce the HRSV burden to public health and better protect the populations at risk.

## Introduction

Human respiratory syncytial virus (HRSV) is one of the most common causes of acute lower respiratory tract infection worldwide, particularly in children less than five years of age, with a reported incidence of >33 million cases globally per annum [1]. Although preterm neonates, infants less than one year of age, the elderly, and the immunocompromised are recognised as the cohorts most susceptible to severe HRSV disease, reinfections in older children and adults with mild presentations also occur, placing a significant burden on economic productivity and healthcare systems [2, 3]. Vaccination has been made available and recommended for adults aged ≥60 years [4] and for pregnant women during weeks 32-36 gestation to afford protection to infants during the first six months of life [5] in the United States of America. Additionally, passive immunisation with monoclonal antibodies, designed to prevent the development of severe HRSV disease, is available for prophylactic administration in vulnerable paediatric cohorts, including palivizumab (Synagis) and, since July 2023, the long-acting intramuscular preparation, nirsevimab (Beyfortus) [6, 7], planned to be offered in Ireland from September 2024 [8].

HRSV is taxonomically classified within genus *Orthopneumovirus*, family *Pneumoviridae*, harbouring a non-segmented, single-stranded, negative-sense RNA genome of ∼15.2 kb, encapsidated within an enveloped virion [9]. Among encoded viral proteins, the attachment (G) and fusion (F) major surface glycoproteins mediate cellular binding, entry and infection [10]. Both G and F elicit immune responses; however, F represents the majority of the primary antigenic determinants for virus-neutralising antibodies [11].

HRSV is classified into two major antigenic subgroups: HRSV-A and HRSV-B, distinguished by mono- and polyclonal antibodies [9]. Such subgroups are further characterised into phylogenetically separable lineages according to genetic distances to other lineages [12]. Prior studies have presented conflicting data regarding the clinical severity associated with each HRSV subgroup: nonetheless, these frequently co-circulate in the winter of countries with temperate climates with alternating seasonal patterns of predominance [2, 9].

With a significant burden for public health in terms of both morbidity and mortality, HRSV has been a notifiable disease in Ireland since 2012 [13]. The introduction of non-pharmacological interventions (NPIs) during 2020 and 2021 to interrupt SARS-CoV-2 transmission was anticipated to reduce the circulation of other respiratory viruses, including HRSV. Subsequently, in the 2022/23 and 2023/24 HRSV seasons, a disproportionately high number of HRSV cases was reported in Ireland with large numbers of hospitalisations arising from seasonal HRSV outbreaks; indicating more complex viral transmission dynamics and epidemiology [14, 15].

To clarify the potential effects of NPIs during COVID-19 on HRSV genetic diversity and to establish a genomic epidemiological baseline prior to the implementation of more widespread active and passive immunisation measures, we have analysed the genomic diversity of HRSV obtained from clinical samples collected both before (2015-2019) and after (2022-2024) the COVID-19 pandemic. We have assessed classification based on both the HRSV G gene ectodomain and complete genome sequences to assess viral diversity and shown it to be a robust approach. We provide evidence of loss of genetic lineages, lineage replacement events and a newly-emerging post-pandemic lineage (A.D.1.4) within HRSV-A which circulated widely in Ireland and other European countries in late 2023. These findings highlight the need for robust and rapid pathogen genomics to better inform public health policies with the availability of new vaccines and monoclonal antibody countermeasures.

## Materials and Methods

### Sample and data collection

Clinical samples were collected and sent to the National Virus Reference Laboratory for laboratory diagnosis by molecular methods for patients presenting with symptoms related to HRSV across Ireland between 2015 and 2024. Data were compiled and summarised from publicly available national disease surveillance reports, produced by the Health Protection Surveillance Centre (HPSC, https://www.hpsc.ie/notifiablediseases/annualidstatistics/) and downloaded between October 2017 and May 2024 (https://respiratorydisease-hpscireland.hub.arcgis.com/, downloaded on May 16^th^). Reference genome sequences for the clades were downloaded from HRSV Genotyping Consensus Consortium repository (https://github.com/rsv-lineages) for HRSV-A (n=246) and -B (n=145) (accession numbers listed in Supplementary Table 1) [12].

### Sample extraction and storage

Nasopharyngeal samples were extracted using the Roche MagNA Pure 96 DNA and Viral NA Small Volume kit according to the manufacturer’s instructions and eluates stored at −80□ prior to use.

### Molecular diagnostics

Respiratory samples received for routine screening were tested using the NxTAG Respiratory Pathogen Panel + SARS-CoV-2 assay (Luminex), which identifies both HRSV-A and -B subgroups. Additionally, samples referred to the NVRL for further characterisation were tested using a lab-developed qPCR, based on protocols described previously [16, 17], which again allowed discrimination between HRSV-A and -B. Specimens with a viral titre yielding results of MDD ≥300 (NxTAG) or Ct ≤25 (qPCR) were considered eligible for sequence analysis.

### HRSV G gene sequencing

Laboratory-confirmed HRSV positive samples from the period 2015-2019 were sequenced by standard Sanger approaches with nested PCR targeting the genomic region corresponding to the G protein ectodomain, from amino acid 78 to the end of the protein [18], using primer sequences provided in Supplementary Table 3, covering 756 out of 966 nucleotides of the gene. Sequences were deposited in GenBank with accession numbers: PP957934-PP958215 and metadata for these samples is provided in Supplementary Table 3.

### HRSV whole-genome sequencing

The reference genome alignment for a diverse group of target sequences was compiled from available complete and nearly complete genomes for HRSV-A and -B in GenBank. Sequences were organized into clusters with *usearch* software with the *cluster_fast* option after sorting by length (http://www.drive5.com/usearch/). Clustering at 2% identity was employed and representative sequences were chosen for each cluster to avoid overrepresentation of common strains or underrepresentation of genetic outliers. Cluster sizes were tracked with the *sizeout* option, resulting on eight clusters for HRSV-A of 461, 243, 114, 85, 62, 42, 15, and 7 sequences, represented by MW582528, KJ643561, KU950673, FJ948820, KJ643484, MG642060, MW768128, and GU591767, respectively. Analogously for HRSV-B yielded four clusters of 810, 75, 15 and 5 sequences, represented by KP317952, KF893260, KP317923, and MN167850, respectively. Cluster-representative sequences were aligned for HRSV-A and -B and used as input in *primalscheme* (https://primalscheme.com/) to design two sets of primers for tiling amplicon multiplex PCR. Resulting primers were aligned to the alignment of all representative cluster sequences with MAFFT [19] using the *addfragments* option, to allow inspection. Alternative primers were added at positions where too many differences were found with the primer-binding sites in sequences representative of the smaller clusters and for missing fragments, based on the sequence of the overlapping part of neighboring fragments. Laboratory-confirmed HRSV positive samples received and stored between 2021 and 2024 were processed for whole-genome sequencing (WGS) with a next-generation sequencing approach employing the RSV subgroup-specific oligonucleotide primer pools (Metabion, Germany) described in Supplementary Tables 2 and 3 upstream of an Illumina DNA library preparation as described downstream of the tagmentation step [20] with a Illumina NextSeq 1000 with 2×150 runs. Assembly of the genomes was performed using the InsaFlu platform (https://insaflu.insa.pt/) that assembles consensus sequences by reference-mapping of the reads, with reference for HRSV-A and -B as EPI_ISL_412866 and EPI_ISL_1653999, respectively. Sequences were made publicly available at GenBank (PP969941-PP970173) and also in GISAID, see accession numbers and reference coverage details in Supplementary Table 4.

### Phylogenetic analysis

Multiple sequence alignments were performed with MAFFT [19]. Phylogenetic trees corresponding to multiple sequence alignments of datasets with (i) G gene and (ii) genome sequences were inferred by maximum-likelihood approaches with IQ-TREE version 2.3.1 [21] and the support for the topology tested with SH-like approximate likelihood ratio test (SH-aLRT) and ultra-fast bootstrap (UFBOOT) with 10^4^ replicates and considering support for branches with values >80% and >95%, respectively. The lineages of sequences were assigned with Nextclade (https://clades.nextstrain.org/) using EPI_ISL_412866 (GISAID) and OP975389 (GenBank) as references for HRSV-A and -B, respectively.

### Phylogenetic compatibility matrix

The divergence across different HRSV genomic regions and its impact on the phylogenetic relationships among sequences was assessed in a sliding window fashion, with window lengths of 500 nucleotides (nt) and step 100 nt. Pairwise evolutionary distance was estimated for each window under a Tamura–Nei evolutionary model, and used to infer a neighbour-joining phylogenetic tree for comparison against trees corresponding to other windows in a compatibility matrix, similar to the approach described previously by Carr and colleagues [22], where the compatibility of two windows is defined as the normalised Robinson–Foulds distance [23] between the corresponding trees. Thus, the compatibility reflected how similar are the inferred phylogenies for any two genome windows ranging from 1 (identical) to 0 (completely dissimilar).

### Graphical and Statistical analysis

Data analysis and graphic representations were performed with R packages and scripts available upon request. χ^2^ tests were used to compare frequencies of lineages per year.

## Results

### HRSV cases in Ireland in 2017-2024

Publicly-available epidemiological data of total HRSV laboratory-confirmed cases reported weekly between October 2017 and May 2024 (n=344 weeks), reflects a sharp increment in the last two HRSV seasons (Fig. 1A). Reported cases reflected the most affected groups, prior to and after the COVID-19 pandemic, corresponding to children under 14 years and adults ≥65 years, consistent with prior reports [1]. Furthermore, the combination of these age-groups declined from 90% (2856/3148) of cases in the season 2017-2018 to 87% (6825/7825) in the season 2023-2024. The HRSV epidemic season is defined in Ireland between week 40 and week 10, i.e. from October to the second week of March of the following year; however, we note there is a tendency to have a notable number of cases accumulating before the season starts (Fig. 1B) in HRSV seasons 2021-2022 and 2022-2023.

**Figure 1.**
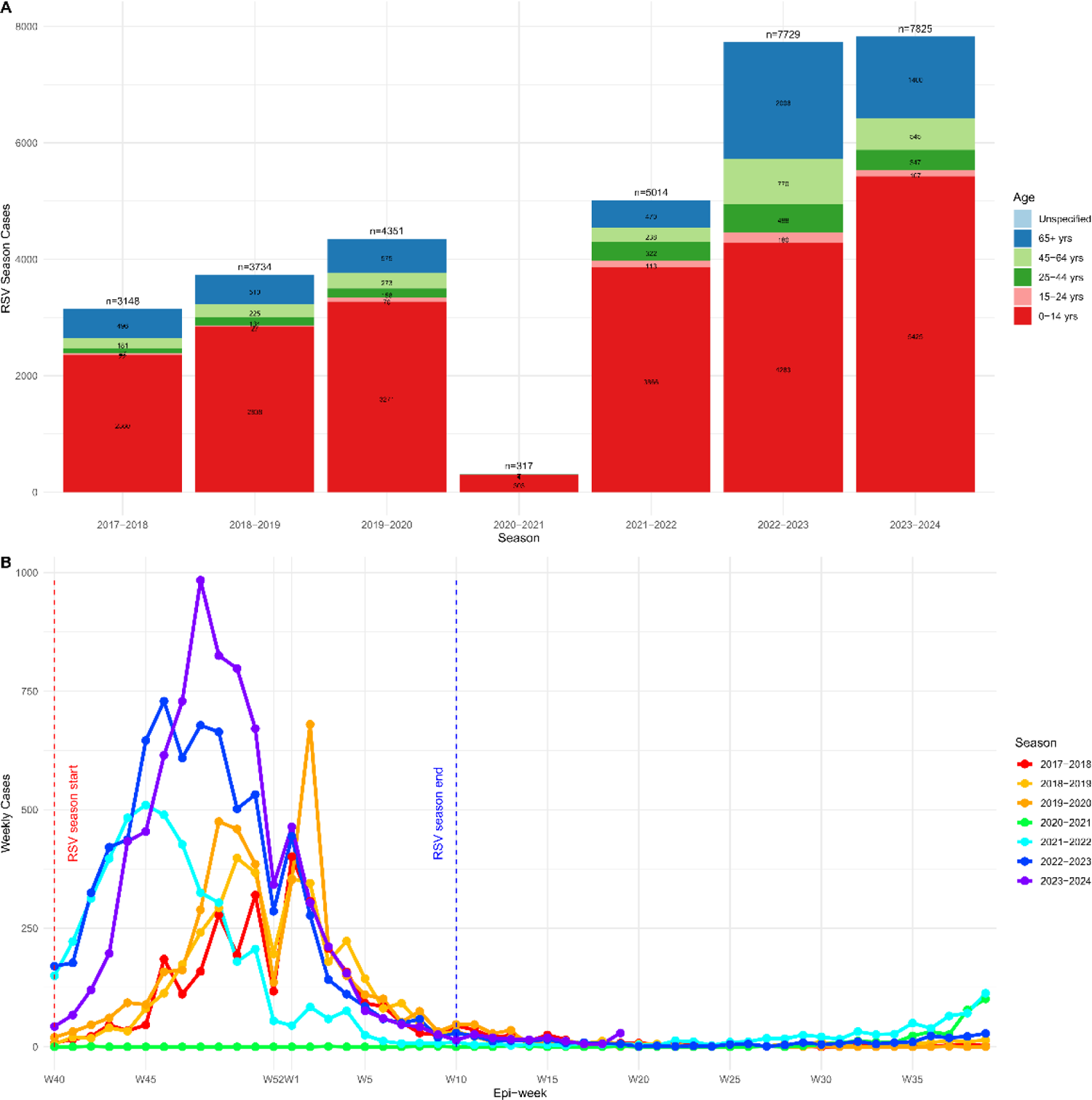
HRSV cases reported between October 2017 and May 2024. **(A)** shows the total number of HRSV cases (vertical axis) reported in Ireland per HRSV season (horizontal axis). The colours for the age ranges of the patients are shown in the legend to the right of the panel. **(B)** shows the details of weekly counts (vertical axis) collected by surveillance programs per week (horizontal axis). The start (red dotted line) and end (blue dotted line) of the HRSV seasons in Ireland are highlighted. The HRSV seasonal series of cases per week are coloured as the legend on the right.

### Assessment of lineage-informative loci in HRSV genomes

We assessed the evolutionary distance among HRSV reference genome sequences of both viral subgroups using a sliding window approach (Fig. 2A-B). The highest nucleotide sequence diversity among sequences was located in the G gene as 0.057 ± 0.042 and 0.045 ± 0.030 for subgroups A and B, respectively (Fig. 2A-B). The G-gene had lower phylogenetic compatibility to other genomic regions (Fig. 2C-D) and to the phylogenetic tree of the whole genome (Fig. 2E-F). The locus encoding the G gene is suggested to be less constrained by structural and functional interactions potentially attributable to humoral selective pressures, therefore represents an informative sequencing target for tracking HRSV lineages and their epidemiological distribution (Fig. 2C-D). It is noteworthy that despite the epidemiologically informative nucleotide variability of the G gene, G is not the target for HRSV prophylaxis, thus limiting the interpretation of surveillance based solely on this gene. G gene evidenced an average pairwise distance among reference sequences of 0.05 ± 0.04 and 0.04 ± 0.03 in the HRSV-A and -B datasets, respectively, while the average pairwise distance at genome sequence level was 0.02 ± 0.02 and 0.01 ± 0.01.

**Figure 2.**
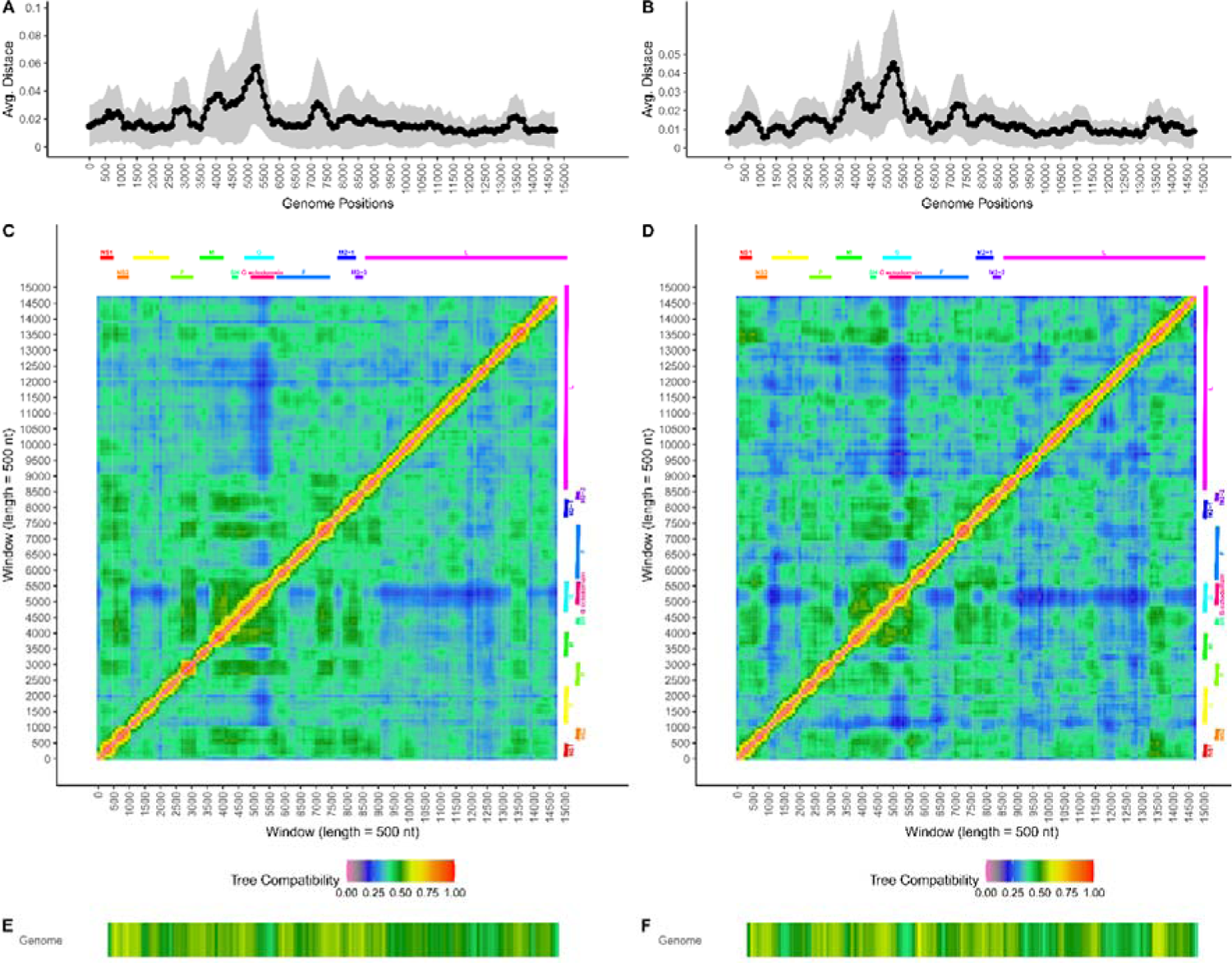
Distribution of genomic diversity along the HRSV-A and HRSV-B genomes. The visualised data corresponds to windows (length = 500 nt) from sliding-window analyses along the multiple sequence aligned reference genomes of HRSV-A and HRSV-B. The horizontal axes represent the genome positions of the windows. **A)** and **B)** present the average evolutionary distance (under a TN93 model) per window represented by the black series. The vertical axis represents the evolutionary distance. The grey shadow shows one standard deviation above and under the average for the windows. **C)** and **D)** Phylogenetic compatibility matrices comparing the compatibility between phylogenetic trees corresponding to the windows of the genome. The vertical and horizontal axes represent the genome positions of the windows being compared. The compatibility is coloured according to the legend at the bottom of the panel. The gene annotation corresponding to the windows is shown at the top and right of the heatmap, including G ectodomain. **E)** and **F)** Phylogenetic compatibility of each window tree against the tree of the corresponding whole genome coloured following the legend on top of the panel.

The virus classification consistency was assessed by comparing lineages assigned using the G locus against lineages assigned using whole-genome sequences of the reference dataset. This showed 96% (235/246) and 99% (144/145) consistency in the assigned lineages to each sample based on G ectodomain and complete genome sequences of HRSV-A and -B, respectively. Among the inconsistencies there were nine HRSV-A reference sequences and only one HRSV-B misclassified (details are shown in Supplementary Table 1). Although phylogenetic trees inferred using the G target and complete genome sequences showed different phylogenetic positions between lineages (Fig. 3A-C), average intra-lineage evolutionary distances confirmed high similarity of sequences under the same lineage and higher differences to sequences in other lineages (Fig. 3B-D). Moreover, the G locus was shown to be a robust target for sequence classification considering the ectodomain region (Supplementary Fig. 1A-B), the tree distance to the genome phylogenetic tree (Supplementary Fig. 1C-D), the shared phylogenetic information (Supplementary Fig. 1E-F), and the mutual clustering information with the complete genome sequences (Supplementary Fig. 1G-H). Despite the relatively shorter sequence length of G locus to the entire genes for L, G and F proteins in HRSV-A and HRSV-B, the G target analysed here presented comparable consistency of lineage classification to those other loci. Therefore, we infer that the lineage classification achieved by the G ectodomain is comparable to the lineage classification based on the whole-genome sequences.

**Figure 3.**
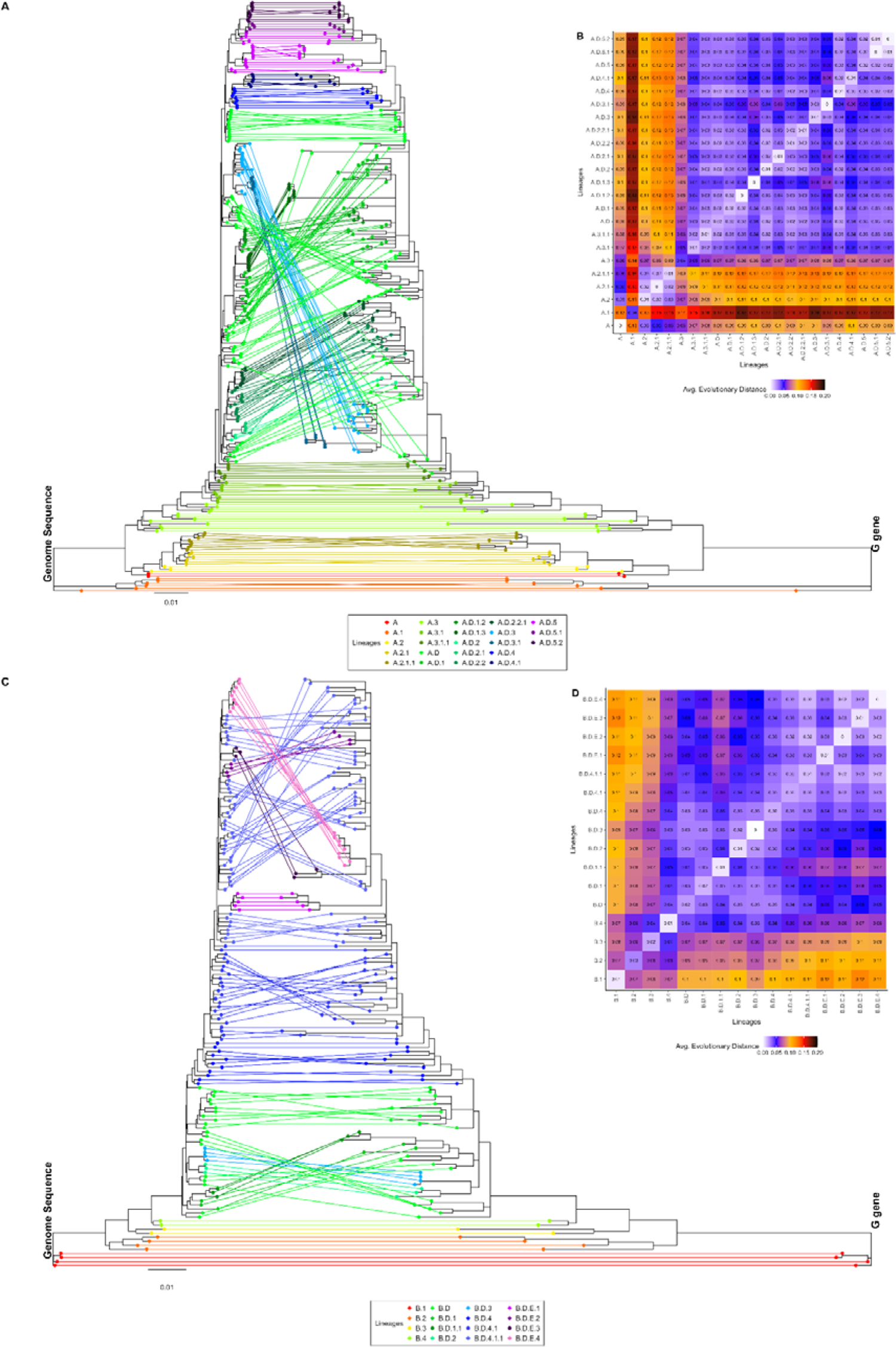
Comparison of tree topologies for HRSV reference sequences. **In panels** (A) and **(C)** the left-hand tree shows the phylogenetic tree inferred for the complete genome sequences of **(A)** HRSV-A and **(B)** HRSV-B and the right-hand phylogenetic tree inferred for the corresponding G target sequences. Sequences in both trees and lines connecting them are coloured according to the lineages as shown in the legend at the bottom of each panel. Panels (C) and (D) show the average evolutionary distance between G gene sequences of clades showed in the horizontal and vertical axes.

### Epidemiology of HRSV cases in 2015-2024 in Ireland and Europe

The available Irish HRSV lineage data in 2015-19 corresponds to lineages assigned employing G gene sequencing. The lineages from 2015-2019 were compared against lineages detected in Ireland in 2022-2024 and assigned with complete genome sequences for both viral subgroups (Fig. 4). In HRSV-A, clusters of samples in years 2022 and 2023 indicated decreased viral lineage diversity (Fig. 4A). Furthermore, HRSV-B lineage analysis demonstrated the prevalence of a single lineage during the same period (Fig. 4B). Nevertheless, comparisons of pairwise evolutionary distances between the different sampling years supported a significantly greater evolutionary diversity in the G locus for both groups in samples from 2022 and 2024 than in samples previous to 2020 (p < 0.05, χ^2^ test) (Supplementary Figure 2) with the exception of HRSV-B in 2016, which showed a the most diverse range of genetic lineages (Fig. 4).

**Figure 4.**
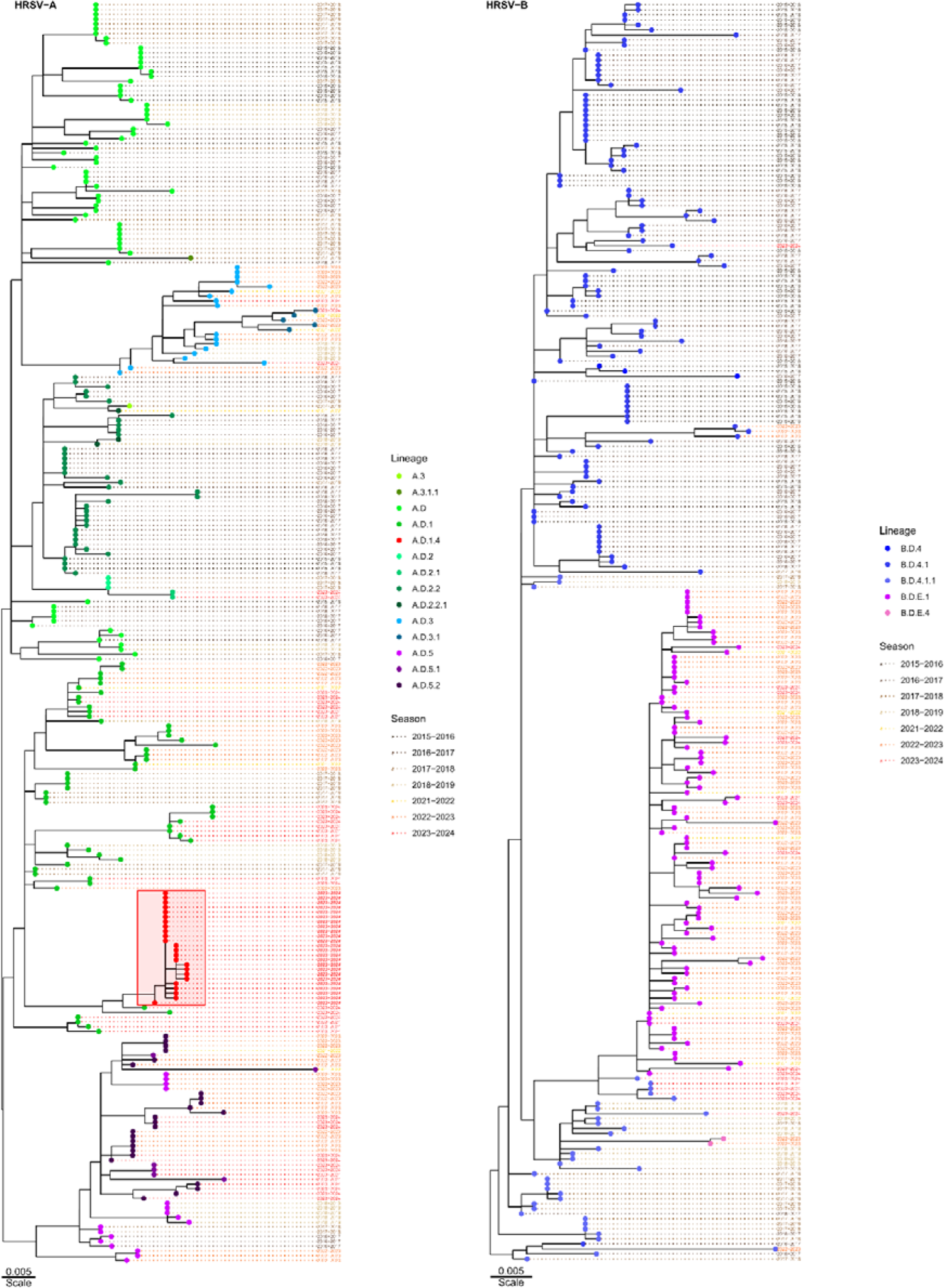
Phylogenetic trees of HRSV cases in Ireland between 2015 and 2024. The tips are coloured according to the assigned lineages shown on the right panels for HRSV-A and HRSV-B. Tip names show the HRSV season. The scales represent the branch lengths in base substitutions per site. Tips of samples under the new lineage A.D.1.4 are highlighted within a red box in HRSV-A.

The predominant lineages per year were further explored for HRSV-A (Fig. 5A) and HRSV-B (Fig. 5B). In both subgroups, we found statistically significant changes in the frequencies of predominant lineages between years preceding the COVID-19 pandemic (2015-2019) and the first years after the NPIs introduced during the pandemic were lifted (2022-2024). In HRSV-A, significant changes in the number of cases per lineage were seen (*p* < 3×10^-33^, χ^2^ test). The first period was dominated by lineages A.D, A.D.2.2 and in lesser degree A.D.1; in the second period, the predominance changed to A.D.1 with a larger percentage (32%, n=123) of the cases, A.D.5 phylogenetically derived lineages, and a novel lineage A.D.4.1 representing 20% of cases. Similarly, the HRSV-B showed a significant predominance change (*p* < 9×10^-45^, χ^2^ test) from B.D.4.1 and later B.D.4.1.1 predominant in the pre-pandemic period, to a predominance of B.D.E.1 after the pandemic.

**Figure 5.**
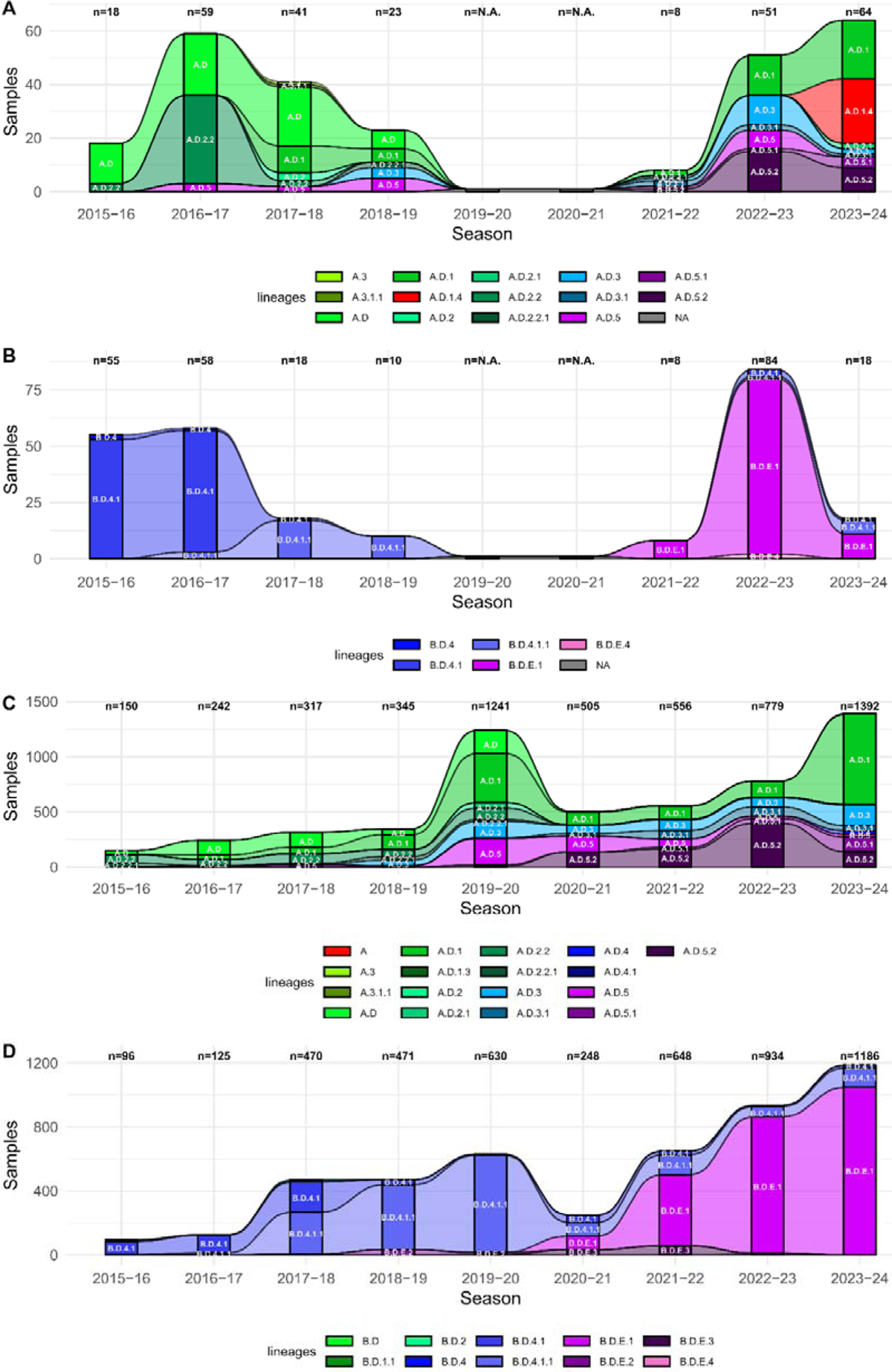
HRSV clades detected in Ireland and Europe between 2015 and 2024. The total number of cases sequenced (vertical axes) per HRSV season (horizontal axes) and stratified and coloured per detected clades as shown in the legend at the bottom of each panel. **(A)** and **(B)** show the data for HRSV-A and HRSV-B, respectively, in Ireland, while **(C)** and **(D)** show the data for HRSV-A and HRSV-B in the rest of Europe. Clades with >1% of the yearly sequences are shown in white letters in the respective stratum.

The changes in the lineage predominance circulating prior to and following the COVID-19 pandemic indicated the replacement of circulating viral genetic lineages in Ireland. To assess this interpretation, lineages circulating in Europe (excluding Ireland) were compared over the same time period, by analysing the whole-genome sequences reported in GISAID for both HRSV-A (Fig. 5C) and HRSV-B (Fig. 5D; Supplementary Table 4). Notably, this separate international analysis also yielded significant results for both subgroups when comparing similar periods 2015-2019 and 2022-2024 (*p* < 9×10^-100^, χ^2^ test).

The higher number of sequences in the European dataset contextualised the observations from the national dataset reflecting very similar trends, with an increase in the prevalence of A.D.1 (44%, n=961/2171) and A.D.5.2 (25%, n=536/2171) for the HRSV-A subgroup, and a change in prevalence of B.D.E.1 in HRSV-B in recent years (90%, n=1900/2120).

### Novel HRSV-A lineage circulating in Ireland and internationally

Our genomic surveillance allowed the detection and characterisation of a novel HRSV-A lineage descendant from A.D.1 (Fig. 4A) under the standardised nomenclature criteria recently established by Goya and co-workers [12]: must present ≥five amino acid mutations distributed among multiple proteins and form a monophyletic, highly-supported cluster of multiple sequences. We identified 24 HRSV-A sequences classified within the A.D.1 lineage with at least eight characteristic amino acid substitutions (Fig. 6A): three substitutions in the G protein, one substitution in the F protein, and four substitutions in the L protein. The three substitutions in G protein, A122V, I133V and Y273H, reside in the two mucin-like domains (domain I: 66-164 and domain II: 198-298)[18], which could potentially impact immune evasion. Substitution in F protein, L3S, located in the N-terminal domain could have effects on protein processing as the section is proteolytically removed in the mature F protein. Finally, four amino acid substitutions in L protein are located in multiple domains associated with virus replication and infection: notably, D146G in the RNA-dependent RNA-polymerase (RdRp) domain; I1656T and G1735D are located in the connector domain between capping and methyltransferase domains, and V1934I located in the mononegavirus-type SAM-dependent 2’-O-MTAse. Assessing the impact of these substitutions requires further experimental characterisation. This novel lineage has been formally named A.D.1.4 following the current nomenclature and procedures for assignment of a new viral genetic lineage. These substitutions as criteria allowed identifying other 90 instances of this lineage circulating in Europe (n=53), Australia (n=12), Brazil (n=15) and USA (n=10) (Supplementary Table 5), with 63 of those preceding cases in Ireland.

**Figure 6.**
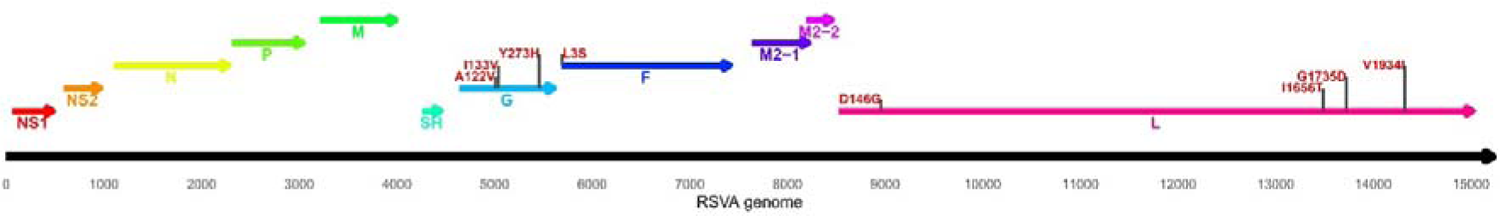
Amino acid substitutions present in the novel HRSV-A A.D.1.4 lineage. Annotated HRSV-A genome representation with the distribution of genes and amino acid substitutions distinguishable from other lineages. The horizontal axis corresponds to genome positions the predicted products are annotated as arrows. Substitutions positions are annotated with vertical black lines and the amino acid changes in red, indicating the reference amino acid, position relative to the protein and the substitution amino acid.

### Impact of diversity on available treatments

The amino acid conservation of F protein among all Irish samples was analysed in 2022-2024 to identify substitutions in the binding sites to the monoclonal antibody nirsevimab in residues 62-69 and 196-212, following results reported by Wilkins et al. [24]. Among HRSV-A, 3 out of 124 (2%) sequences showed a substitution affecting site 65 in the F2 subunit from lysine (K) to arginine (K65R) in clade A.D.3.1, which *in vitro* has been reported to result in a <10% fold-change in IC_50_ [24]. HRSV-B exhibited greater divergence in F1 subunit sites, with isoleucine (I) at residue 206 substituted by methionine (M) in 107 out of 111 sequences (I206M), consistent with the observation of this substitution being more prevalent in recent samples and in vitro leading to a <10% fold-change in IC_50_ [24]. Moreover, sites 209 and 211 also showed substitutions from arginine (R) and asparagine (N), respectively, to glutamine (Q) (5 out of 111 sequences) and serine (S) (8 out of 111 sequences) with these substitutions concurrent in four sequences of B.D.4.1, one B.D.E.1 showing the R209Q, two B.D.E.4 and one B.D.4.1.1 lineages showing the N211S substitution. These substitutions were recently identified and require further characterisation to assess any potential impacts on prophylactic efficacy. We performed a similar analysis focused in the palivizumab neutralising epitope in F amino acid residues 258-275, following the results reported by Hashimoto and Hosoya [25], however, no substitutions were found affecting this region of F protein in either viral subgroup.

## Discussion

Our genomic surveillance revealed that COVID-19 containment measures disrupted the HRSV seasonality during the 2020-2021 season in Ireland, suggesting these interventions provided protective effects against the spread of HRSV and other common respiratory viruses. However, HRSV seasons 2022-2023 and 2023-2024 showed increases in cases, likely due to a growing immunologically-susceptible population, waning immunity and enhanced testing capabilities of respiratory infections – potentially augmented by changes in the healthcare-seeking behaviour in younger populations after the COVID-19 pandemic. These periods also showed changes in circulating viral genetic lineages, with marked increases in A.D.5.2 in HRSV-A and B.D.E.1 in RSV-B, reflecting a regional shift in lineage prevalence potentially influenced by a HRSV transmission bottleneck-effect during the COVID-19 pandemic. Additionally, we also identified a novel post-pandemic HRSV-A lineage circulating in Ireland and internationally since 2023. Importantly, we have detected HRSV-B strains characterized by two notable substitutions in F protein (R209Q and N211S) which may affect the efficacy of the long-acting monoclonal antibody preparation, nirsevimab.

The current study provides evidence supporting the comparability of HRSV clade classification data based on partial sequencing of a 756-nucleotide segment of the G gene against complete genome sequences in both subgroups, HRSV-A and HRSV-B, consistent with recent findings by Goya and colleagues [26]. Such evidence enabled the comparison and epidemiological analysis of HRSV genetic lineages circulating in Ireland during 2015-2019 against lineages circulating in 2022-2024 following the lifting of NPIs introduced to prevent SARS-CoV-2 transmission.

Although our results support the informative role of sequencing the G gene for clade classification and surveillance information, they also demonstrate the value of more comprehensive whole-genome sequencing methods to assess viral divergence across the entirety of the HRSV genome, particularly within the vaccine-relevant F external glycoprotein to assess changes in the frequency of amino acid substitutions as pharmaceutical countermeasures become available. Molecular surveillance also enables computational analysis of the PCR-based molecular diagnostic testing, characterisation of seasonal epidemics and the efficacy of on-going responses and putative therapies. Such efforts are similar to current influenza surveillance programmes. We identified two substitutions in the F protein of HRSV-B that could affect the efficacy of treatments with nirsevimab, R209Q and N211S, which require further functional characterisation to assess their impact.

Also, our genomic surveillance initiative allowed the characterisation of a novel lineage circulating in Ireland during 2023 named A.D.1.4. This lineage, descendant from A.D.1, is related to sequences from a number of other European countries exhibiting the same distinguishable substitutions since 2020. Although the effects of the reported substitutions remain to be experimentally explored, the growing frequency across international borders of a lineage that appeared during the COVID-19 pandemic is consistent with our observations of changes in the HRSV lineage dominance with possible relation to the impact of NPIs on extant viral genetic diversity. These changes in lineage abundance, lineage replacement events and the emergence of new genetic lineages necessitate the continuous revision and updating of the WGS primer pool approaches and the lineage designation to properly quantify and track epidemiological trends and potential changes in risk identification, as also suggested by Goya [12].

We were limited by the number of samples with enough recoverable genetic material to be sequenced despite the number of yearly cases limiting the sequencing rate to 1-2% of Irish cases. Cases spanning the period 2020-2021 were not stored for sequencing as the workload demands and focus on SARS-CoV-2 precluded the storage and subsequent analysis of HRSV lineage distribution during this period. This limitation was partially addressed by analysing the lineages circulating in the European region.

In conclusion, this study represents the first comprehensive analysis of HRSV lineages circulating in Ireland, spanning pre- and post-COVID-19 pandemic periods demonstrating notable shifts in viral genomic diversity likely attributable to the widespread institution of NPIs between 2020 and 2021. Our data also offer an important baseline for future analyses of HRSV genomic diversity in the context of anticipated novel preventive modalities, and a framework to monitor for the emergence of variants with potential to impact treatment efficacy, or vaccine escape.

## Ethical statement

This study analyses publicly available data collected during the surveillance and report of HRSV in Ireland. The collection analysis of this data was reviewed by the Human Research Ethics Committee Sciences of the University College Dublin with the reference number LS-LR-23-226-Carr.

## Data Availability

All data produced in the present work are contained in the manuscript or is part of the supplementary data available online: https://doi.org/10.5281/zenodo.12797565

## Acknowledgements

We wish to thank all laboratory staff and consultant microbiologists at the referring laboratories for forwarding HRSV positive samples to the NVRL for genetic characterisation, and all Public Health doctors and epidemiologists involved in the surveillance of HRSV in Ireland. The institutions of the following co-authors are partners in Preparing for RSV Immunization and Surveillance in Europe (PROMISE); LP, LA, HV, AM (RIVM). This project has received funding from the Innovative Medicines Initiative 2 Joint Undertaking under grant agreement No 101034339. This Joint Undertaking receives support from the European Union’s Horizon 2020 research and innovation programme and EFPIA. This study was partially supported by AMED under the grant number JP223fa627005. We gratefully acknowledge all data contributors, i.e., the Authors and their originating laboratories responsible for obtaining the specimens, and their Submitting laboratories for generating the genetic sequence and metadata and sharing via the GISAID Initiative and publicly deposited in GenBank, on which this research is based (Supplementary Table 1).

## Author’s Contributions

**Alan Rice**: data analysis and manuscript preparation; **Gabriel Gonzalez**: data analysis and manuscript preparation; **Michael Carr**: genome sequencing and manuscript preparation; **Jonathan Dean**: data collection and analysis, manuscript preparation; **Emer O’Byrne**: sample sequencing and data collection; **Lynn Aarts**: WGS sequencing development; **Harry Vennema**: sequencing protocol design; **Weronika Banka**: sample preparation and data collection; **Charlene Bennett**: data collection; **Siobhán Cleary**: data analysis; **Lisa Domegan**: data analysis and clinical interpretation; **Joan O’Donnell**: data analysis and clinical interpretation; **Maureen O’Leary**: data analysis and clinical interpretation; **Stephanie Goya**: taxonomic validation and interpretation; **Lance Presser**: sequencing protocol and validation; **Adam Meijer**: sequencing protocol and validation; **Greg Martin**: public health interpretation and manuscript edition; **Hirofumi Sawa**: manuscript edition; **Allison Waters**: manuscript edition and data interpretation; **Cillian De Gascun**: results interpretation and manuscript edition; **Daniel Hare**: results interpretation and manuscript edition.

