## Supplementary Materials for "Human Respiratory Syncytial Virus Genetic Diversity and Lineage Replacement in Ireland pre- and post-COVID-19 pandemic"

Supplementary Table 1 to 5 are datasets of metadata publicly available at: <https://doi.org/10.5281/zenodo.12797565>

**Supplementary Table 1**. NextClade Reference Sequences publicly available at <https://github.com/rsv-lineages>.

**Supplementary Table 2**. Whole-genome sequencing library preparation protocol template with concentrations of the primers and reagents for amplification and sequencing.

**Supplementary Table 3**. Primers for sequencing protocols targeting the locus encoding the G protein ectodomain and the whole genome sequencing.

**Supplementary Table 4**. Irish RSV Sequence Details contains details on the sequences generated in our study with the GenBank accession number, the GISAID accession number, reported collection RSV season, Nextclade assigned taxonomical clade.

**Supplementary Table 5**. International RSV Sequences Details contains a list of European RSV sequences downloaded from GISAID on May 2024 (<https://gisaid.org/>) and sequences identified as part of the A.D.4.1 lineage on July 2024, with details on the sampling RSV season, the accession number, country of collection, and lineage assigned by NextClade.

**Supplementary Figures.**

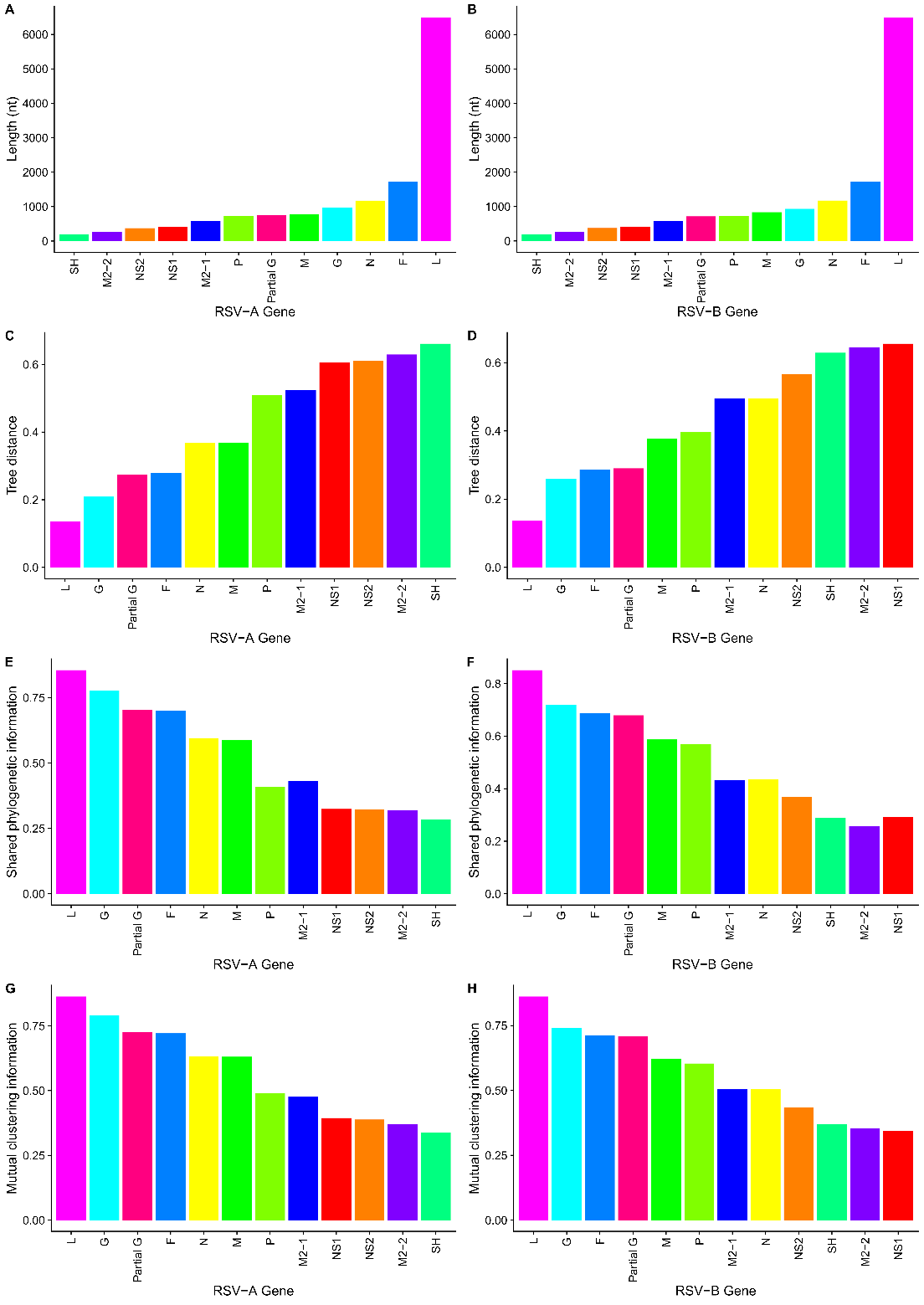

**Supplementary Figure 1. Multiple measures of phylogenetic tree consistency between the whole-genome sequences and HRSV genes. A)** and **B)** show the length in nucleotides of genes and the segment partial G sorted from shortest to largest. **C)** and **D)** show the tree distance to the phylogenetic tree inferred with the complete genome sequences sorted from most similar to most dissimilar. **E)** and **F)** show the shared phylogenetic information sorted from the gene with the most shared phylogenetic information to the least. **G)** and **H)** show the mutual clustering information with the most mutual clustering to the left.

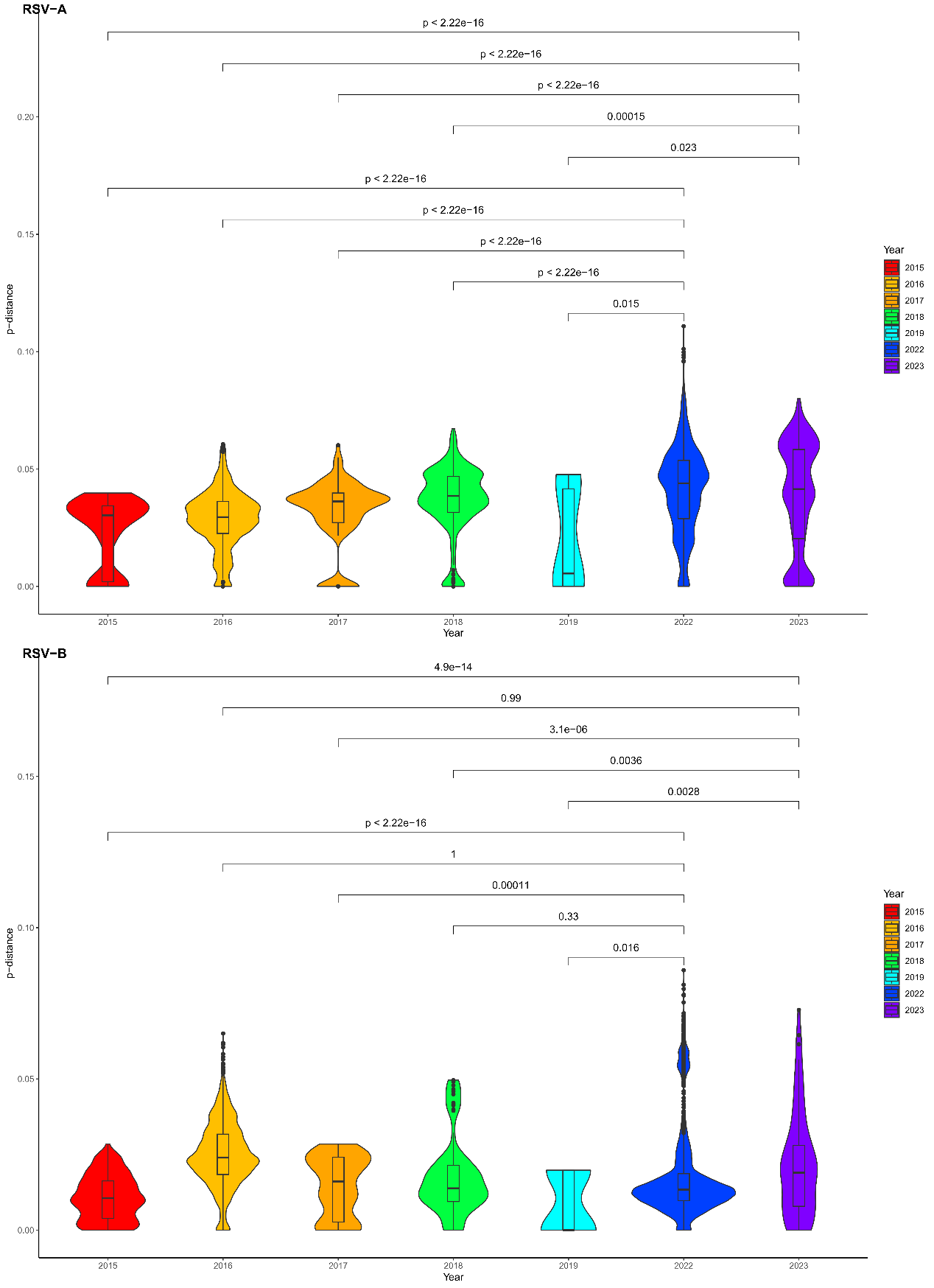

**Supplementary Figure 2. Comparison of average p-distance among sequences per year in the G-target section.** The means of the distributions are compared to test whether the distribution of 2022 or 2023 has higher mean evolutionary distance than the distributions previous to the SARS-CoV-2 2020 pandemic. Distributions are coloured according to the legend on the right of the panels.
